# Evaluating the Efficacy of Large Language Models for Systematic Review and Meta-Analysis Screening

**DOI:** 10.1101/2024.06.03.24308405

**Authors:** Ronald Luo, Ziya Sastimoglu, Abu Ilius Faisal, M. Jamal Deen

## Abstract

**Background:** Systematic reviews and meta-analyses are essential for informed research and policymaking, yet they are typically resource-intensive and time-consuming. Recent advances in artificial intelligence and machine learning offer promising opportunities to streamline these processes.

**Objective:** To enhance the efficiency of systematic reviews, we explored the automation of various stages using GPT-3.5 Turbo. We assessed the model’s efficacy and performance by comparing it against three expert-conducted reviews across a comprehensive dataset of 24,534 studies.

**Methods:** The model’s performance was evaluated through a comparison with three expert reviews, utilizing a pseudo-K-folds permutation and a one-tailed ANOVA with an alpha level of 0.05 to ensure statistical validity. Key performance metrics such as accuracy, sensitivity, specificity, predictive values, F1-score, and the Matthews correlation coefficient were analyzed using two sets of prompts.

**Results:** Our approach significantly streamlined the systematic review process, which typically takes a year, reducing it to a few hours without sacrificing quality. In the initial screening phase, accuracy, specificity, and negative predictive values ranged between 80% and 95%. Sensitivity improved markedly during the second screening phase, demonstrating the model’s robustness when provided with more extensive data.

**Conclusion:** While ongoing refinements are needed, this tool represents a significant advancement in research methodologies, potentially making systematic reviews more accessible to a wider range of researchers.

**Impact Statement:** Our manuscript presents a novel review screening protocol built using open-source frameworks, which significantly enhances the systematic review process in terms of efficiency and cost-effectiveness. Leveraging the capabilities of GPT and embedding models, our protocol demonstrates the potential to transform a traditionally time-consuming and expensive task into an accelerated and economical operation, all while maintaining high standards of accuracy and reliability.

**Key Points:**

- GPT screening can streamline systematic reviews from a year-long, expensive process to just hours at minimal cost.
- Validated across different topics, the protocol exhibits high reliability and consistency in study inclusion.
- The AI-driven process reduces human bias, with prompt optimization considerably improving sensitivity.

## 1. Introduction

### 1.1 Current Challenges of Conducting a Review

Systematic reviews and meta-analyses are cornerstone methodologies in evidence-based medicine, providing a comprehensive synthesis of research findings to inform clinical and policy decisions [1]. However, the traditional approach to conducting these reviews is labor-intensive and time-consuming, often requiring a year or more to complete and significant financial resources [2–5]. The exponential growth in scientific publications further complicates the task, increasing both the complexity and the scope of reviews [6]. This scenario underscores a critical need for innovative methodologies that can streamline the review process without compromising its methodological rigor and accuracy.

The challenges of conducting systematic reviews extend beyond mere resource allocation. The inherent delay in incorporating the latest research findings into reviews due to publication lags adversely affects the timeliness and relevance of the synthesized evidence [6–9]. Additionally, the manual screening process, a key step in reviews, is not only time-consuming but also prone to inconsistencies and biases despite the expertise of reviewers [10–12]. The evolving landscape of systematic reviews, including rapid reviews and evidence synthesis for emergent health issues [6], further demands adaptive and efficient review processes that can cope with the dynamic nature of scientific research.

### 1.2 Text Mining and Automation in Systematic Reviews

The integration of text mining and automated technologies provides valuable tools for overcoming the challenges inherent in systematic reviews. Text mining, a field within data science, involves analyzing unstructured text to extract meaningful information and supports various stages of the review process such as study identification, screening, and data extraction—stages that are traditionally manual and labor-intensive [11–17]. These advancements have the potential to not only speed up the review process but also enhance the accuracy and objectivity of the data extracted, thereby improving the quality of systematic reviews.

Natural Language Processing (NLP), closely related to text mining, enables computers to understand and process human language, playing a crucial role in the automation of systematic reviews. It includes a range of tasks from information retrieval—where relevant articles are identified from large document collections—to document classification, which automates the inclusion or exclusion decisions in systematic reviews [6,18]. With the advent of Large Language Models (LLMs) such as GPT [19], BERT [20], and their successors [19,21–25], NLP has seen significant advancements, offering sophisticated capabilities for text analysis, and understanding that greatly enhance the review process [18]. These technologies allow for more nuanced and comprehensive analysis of scientific literature, leading to more sophisticated and scalable review methodologies.

### 1.3 Role and Impact of AI and LLMs

The integration of Artificial Intelligence (AI) and LLMs into the domain of systematic reviews marks a significant shift towards more efficient and effective evidence synthesis [26]. AI and NLP technologies automate the extraction and analysis of data from vast amounts of literature, streamlining the review process while maintaining, if not enhancing, the depth and breadth of analysis. The role of LLMs, characterized by their large parameter spaces and capacity for unsupervised learning, is particularly noteworthy. These models have demonstrated exceptional ability in understanding context, semantics, and the subtleties of language [27–38], making them well-suited for tasks such as literature screening and data extraction in systematic reviews.

This study focuses on the application of LLMs in the article screening process. By automating this initial screening, AI tools allow researchers to dedicate more time to the complex tasks of data synthesis and interpretation. Moreover, AI-driven processes can potentially enhance consistency and reduce bias by standardizing the application of inclusion and exclusion criteria. As the field progresses, the role of AI and LLMs in systematic reviews is increasingly becoming a cornerstone for enabling more accessible, timely, and rigorous evidence synthesis, which is critical for informing healthcare policy and practice.

## 2. Methods

### 2.1 Overview of Original Reviews

Our research team recently conducted three separate reviews, each varying in degree and scope, and meticulously adhered to protocols established by standardized reporting committees [13,39]. The first review investigated the relationship between dementia and spatiotemporal gait patterns to identify distinctive gait signatures. The second examined the latest advancements in cuffless blood-pressure monitoring devices, while the third focused on the impact of aging and comorbidities on long-COVID [40–42]. The selection process for each review involved pairs of authors screening titles, abstracts, and full texts, with any discrepancies resolved by a third author. Original inclusion and exclusion data were processed and gathered in .csv files, serving as our ground truth for the subsequent comparative analysis and validation depicted in our figures.

### 2.2 Model Selection and Cost Analysis

Since the launch of ChatGPT in November 2022, the landscape of AI and language models has rapidly evolved, offering a range of options for various applications, including systematic reviews. During the planning phase of this study, back-of-the-envelope calculations identified GPT-3.5 Turbo as the most economical model, costing approximately $0.31 USD per 1,000 studies for a single pass of title-abstract screening. In contrast, GPT-4 Turbo was estimated to cost about 21 times more, at $6.61 USD per 1,000 studies. The newest model, GPT-4o, released in May 2024, is expected to cost $3.31 USD per 1,000 studies—about 50% less than GPT-4 Turbo—making it a promising candidate for future analysis.

### 2.3 Recreating Title-Abstract Screening with AI Models

Utilizing the LangChain framework [43], the title and abstract screening process was replicated using the OpenAI model GPT-3.5-turbo-0125, employing Retrieval Augmented Generation (RAG) to accurately reflect the inclusion and exclusion criteria of the original human reviewers. In this RAG setup, the application provided a user interface with a text box where reviewers could enter any prompt. This feature allowed the reviewer to design a single prompt that was applied consistently across all titles and abstracts. For our screening protocol, the model was fed prompts based on the inclusion criteria of each review, designed to elicit binary ‘true’ or ‘false’ responses, simulating the decision-making framework of systematic reviews. The responses, along with associated metadata, were captured, cleaned for accuracy, and then exported as .csv files.

### 2.4 Recreating Full-Text Screening with AI Models

For the full-text screening, we adapted our process to accommodate the OpenAI API’s token limit of 4,096. After converting PDFs of selected studies into text format, we segmented these texts using a text splitter into snippets of 1,000 tokens (approximately 750 words) with an overlap of 100 tokens. Each segmented article was individually stored using Hierarchical Navigable Small World (HNSW) vector storage, which supports efficient spatial searches. This setup employed the OpenAI model, text-embedding-ada-002-v2, functioning similarly to a search engine, that uses cosine similarity to rank all the generated text snippets based on their relevance to a user-generated query.

In the full-text RAG setup, the same inclusion prompts designed to elicit binary ‘true’ and ‘false’ responses were used. Both the prompt and relevant text snippets were combined to form the input for GPT-3.5-turbo-0125. We configured the system to return three relevant snippets per query to effectively manage the constraints imposed by the API’s token limit. This strategy leaves sufficient space (approximately 1,096 tokens) for users to employ complex query templates designed for either study screening or data extraction purposes.

### 2.5 Comparison Criteria and Discrepancy Analysis

To assess the effectiveness of GPT-3.5 Turbo’s screening against the original human reviewers, we employed confusion matrices for both the title-abstract and full-text screening phases. Our comparison utilized key performance metrics such as accuracy, sensitivity, specificity, positive predictive value (PPV), negative predictive value (NPV), F1-score, and the Matthews Correlation Coefficient (MCC).

Discrepancy analysis was conducted by first identifying all true positives, true negatives, and false positives through a comparison of GPT-3.5 Turbo’s decisions against reviewer judgments during the title-abstract screening phase and securing the full texts for these articles. The analysis focused specifically on the subset of articles that were mutually recognized as ‘included’ by both the model and the reviewers at the title-abstract phase, ensuring that our evaluation of the model’s full-text screening was grounded in a directly comparable set of studies. Additionally, we assessed the consistency of the AI model by examining the remaining articles that GPT-3.5 Turbo initially classified as ‘included’ during the title-abstract phase but were later ‘excluded’ or continued to be ‘included’ in the full-text stage.

### 2.6 Evaluation of ChatGPT-3.5 Turbo Performance and Validation Procedure

To assess the performance of the LLM against a standardized control, we established a baseline using random binary classification. In this control setup, studies were arbitrarily classified as ‘included’ or ‘excluded,’ mimicking the binary decision-making process typical in systematic reviews. Alongside this, we conducted a self-validation test with two prompts to assess internal consistency within the LLM’s responses, comparing agreement between the model’s decisions when operating in explanation mode versus non-explanation mode.

To further evaluate our model, we adopted a permutation test similar to K-Folds Cross Validation. This involved shuffling the data and dividing both the model-generated dataset and the control dataset into K equal subsets. We computed standard performance metrics for each partition and then averaged the results across all partitions. Finally, to assess the statistical differences between the model and control, we conducted a One-way Analysis of Variance (ANOVA) comparing the mean performance metrics across the subsets and screening phases, using an alpha level of 0.05 to determine significance.

### 2.7 Data Handling and Ethical Considerations

Our research uses data from studies with established ethical approvals. Full prompt templates are provided as supplementary material. Additional data is available upon request.

## 3. Results

### 3.1 Outcomes of the Original Review

In Table 1, we provide a comprehensive summary of the screening process facilitated by GPT-3.5 Turbo for each of the three reviews. This table includes the number of studies identified, screened, included, and excluded, offering a direct comparison and overview of the model’s performance in the review process.

**Table I:** Overview of Screening Results by Human Reviewers Compared to GPT-3.5 Turbo.

| Criteria | <u>Dementia and Gait</u><br><u>Signatures</u> |  | <u>Cuffless BP</u><br><u>Monitoring</u> |  | <u>Long-Covid</u><br><u>Outcomes</u> |  |
| --- | --- | --- | --- | --- | --- | --- |
|  | Reviewers | Model | Reviewers | Model | Reviewers | Model |
| Studies Screened | 3,245 | 3,245 | 422 | 422 | 20,867 | 20,867 |
| Studies Assessed | 377 | 793 | 202 | 106 | 2,008 | 3,079 |
| Studies Included | 34 | 122 | 25 | 29 | 112 | 137 |
| Studies Excluded | 343 | 158 | 177 | 21 | 1,896 | 103 |

Note: BP – Blood Pressure

### 3.2 Title-Abstract Confusion Matrices for each Review

Figure 1 showcases separate confusion matrices for the title-abstract inclusion task. These matrices illustrate the true positives, false positives, true negatives, and false negatives for GPT-3.5 Turbo in comparison to the original reviewers categorizations.

**Figure 1.**
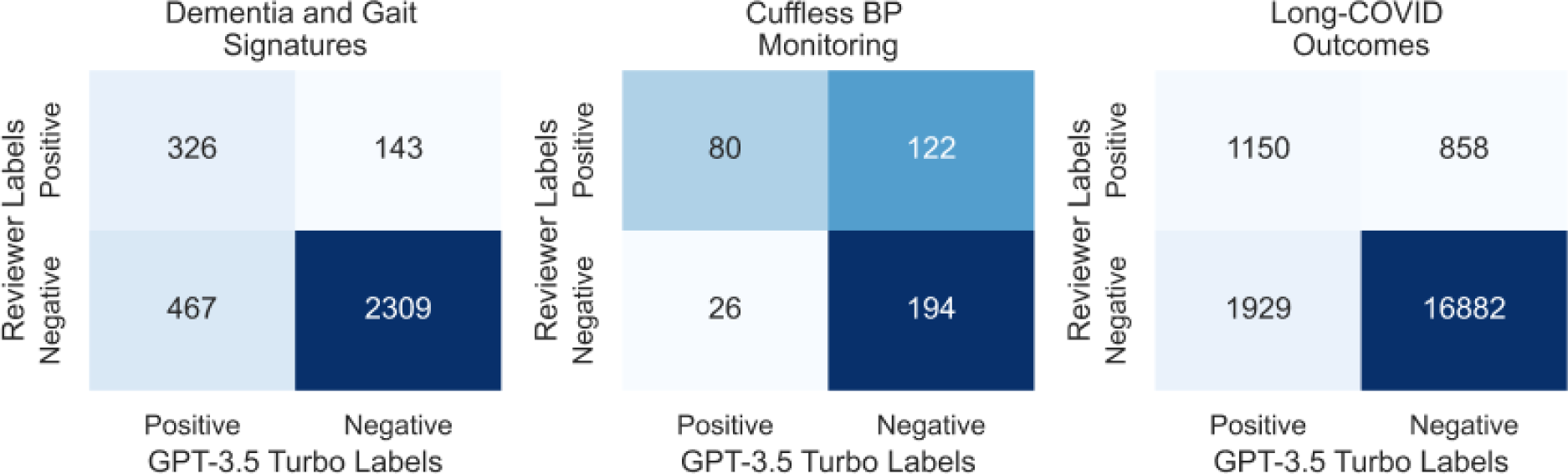
Alignment of Model Predictions with Human Reviewers: Confusion Matrices Comparing GPT-3.5 Turbo Decisions Against Reviewer Judgments for Title-Abstract Screening Tasks in Dementia Gait, Cuffless Blood Pressure Monitoring, and Long-COVID Studies.

### 3.3. Title-Abstract Performance Metrics for each Review

Table II displays the performance metrics of GPT-3.5 Turbo’s responses for each review. This table showcases the model’s accuracy, sensitivity, specificity, predictive values, F1 score, and MCC in comparison to the original human reviewers. It also highlights that GPT-3.5 Turbo significantly outperformed the random classification control in all but two performance metrics (sensitivity: p-value = 0.22; F1-score: p-value = 0.21) emphasizing its effectiveness in accurately identifying relevant studies.

**Table II:**
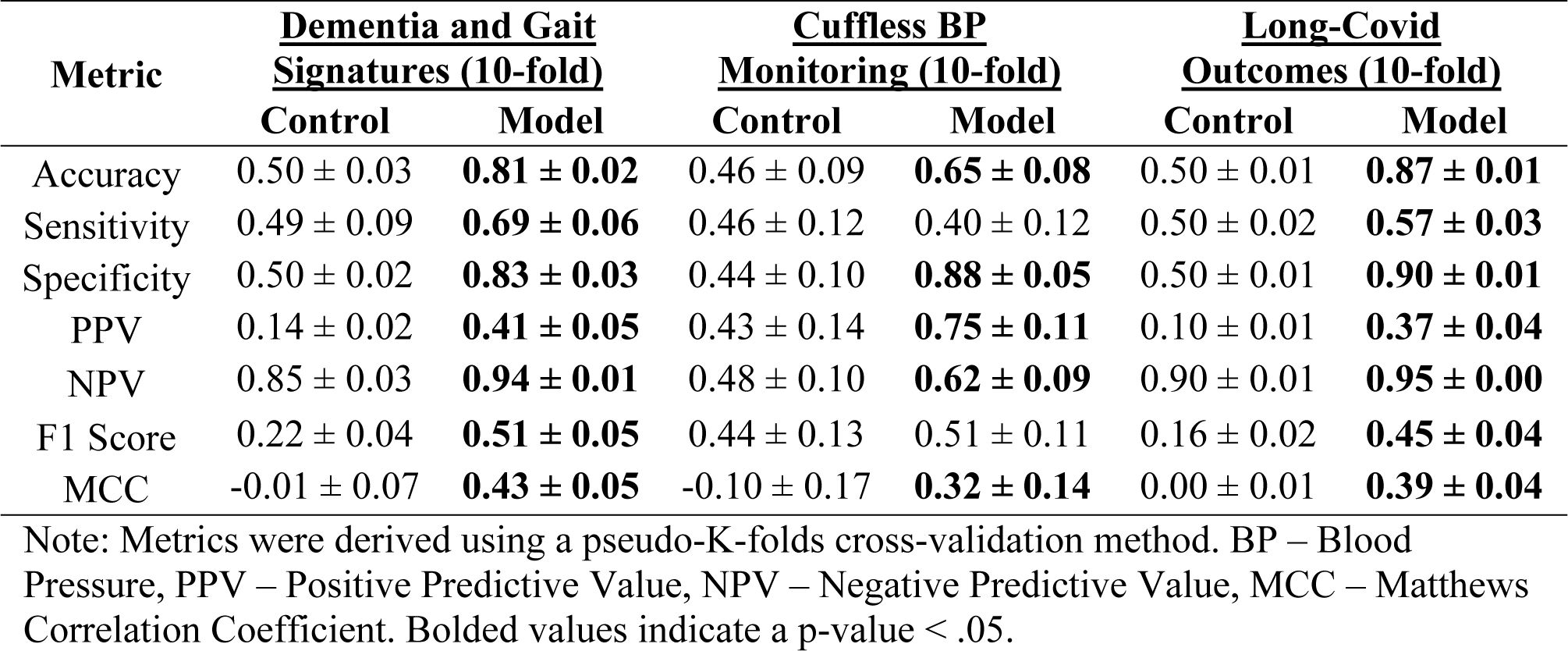
Comparative Analysis of GPT-3.5 Turbo’s Performance Metrics for Title-Abstract Inclusion Across Review Topics.

### 3.4 Title-Abstract Self-Validation of AI Model Responses

Figure 2 presents confusion matrices for the AI model’s responses, comparing when explanations were provided to when they were not. The results show a high level of agreement between the two sets of responses, although not absolute. Notably, when the model’s decisions were matched with those of human reviewers, asking the model for an explanation resulted in a slight drop in performance. This indicates that requesting explanations may subtly affect the model’s output, suggesting the need for further investigation into how such prompts might alter the precision of AI-driven selections.

**Figure 2.**
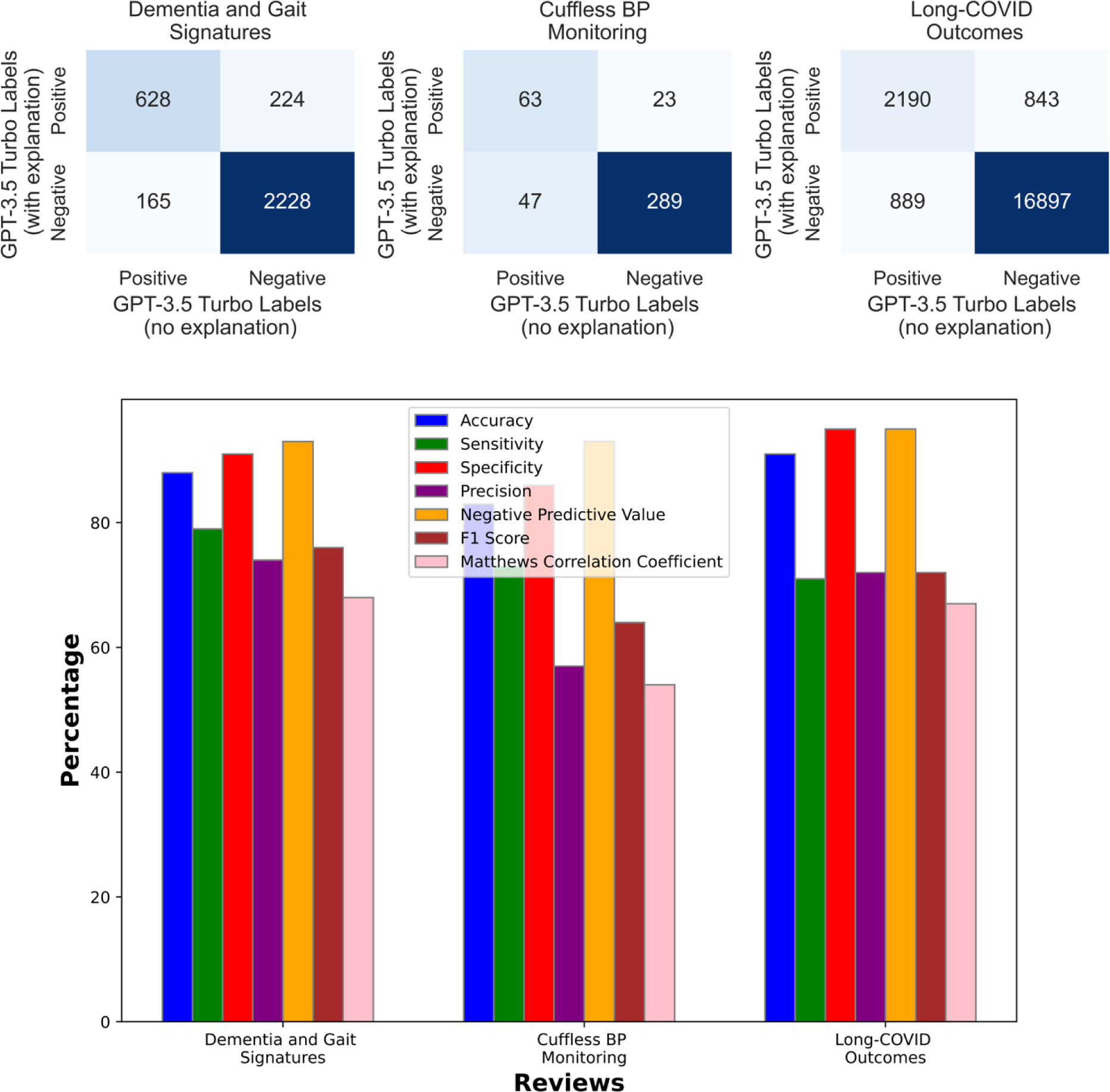
Comparative Analysis of GPT-3.5 Turbo Model Responses: Evaluating Agreement Between Explanatory and Non-Explanatory Assessments Across Dementia Gait, Cuffless Blood Pressure Monitoring, and Long-COVID Outcomes Reviews.

### 3.5 Confusion Matrices for Full-Text Screening

Figure 3 presents the data which compares the AI’s decisions against human reviewers’ judgments. We screened a total of 589 full-text articles on dementia and gait signatures, 64 on cuffless blood pressure monitoring, and 517 on Long-COVID outcomes. The upper section illustrates the alignment of full-text decisions for articles included by both the model and the reviewers in initial phases and the lower section highlights discrepancies for initially included studies.

**Figure 3:**
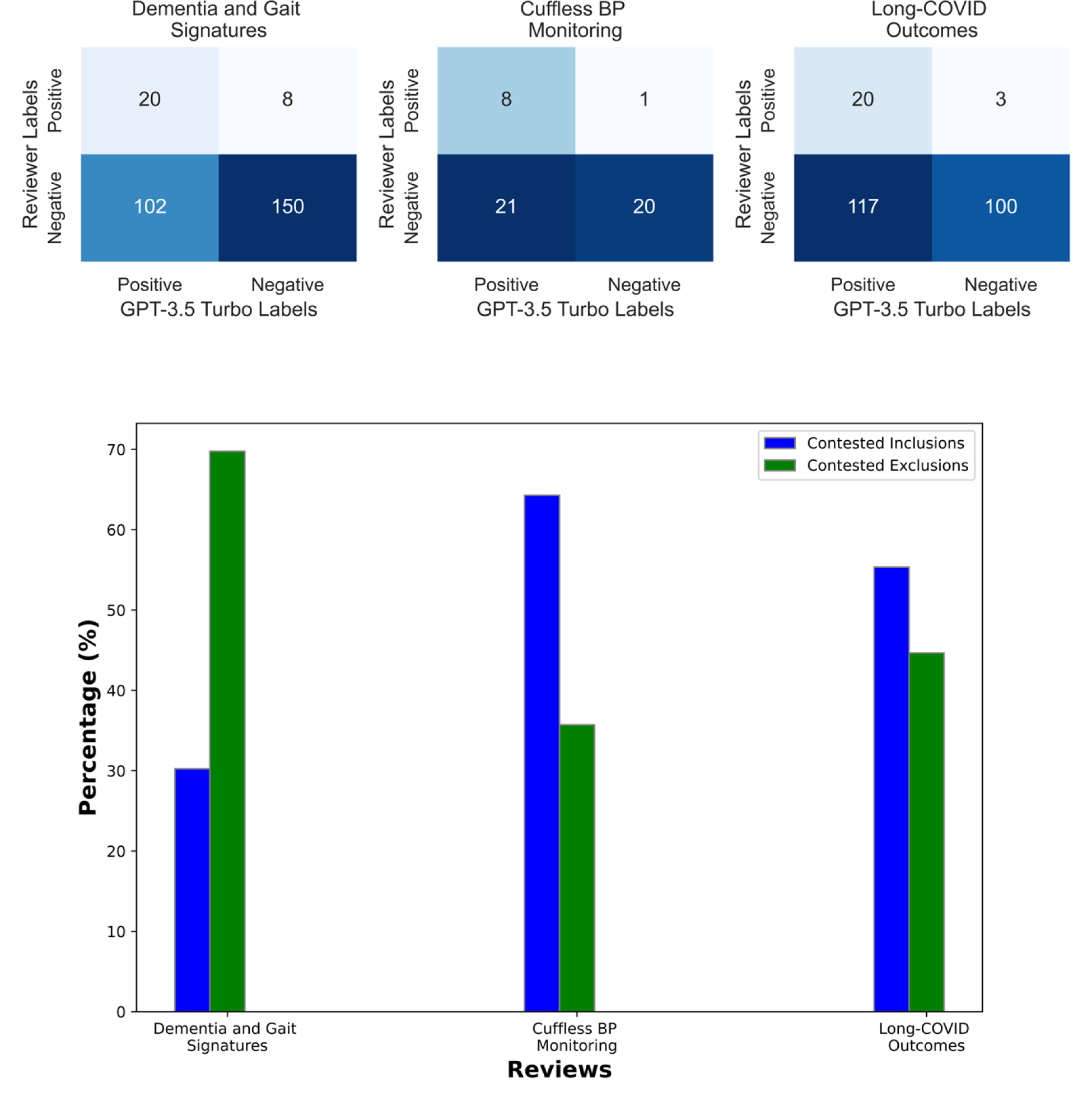
Analysis of Full-Text Inclusion Task and Contested Decisions in AI-Assisted Review Screening. The upper panel presents confusion matrices comparing the AI model’s positive and negative labels against reviewers’ labels. The lower panel (N = 291, N = 14 and N = 262 respectively) shows the percentage of contested decisions differentiating between contested inclusions (blue) and contested exclusions (green).

### 3.6 Performance Metrics for Full-Text Screening

Table III presents the performance metrics of the full-text screening protocol employed by GPT-3.5 Turbo. Sensitivity and MCC improved the most when model screening was applied. Notably, the model had significantly greater sensitivity in the full text screening phase compared to title and abstract screening, indicating a potential reduction in false negatives as more article content is assessed. While the model generally outperformed the control across metrics, many of these improvements did not reach statistical significance possibly due to the smaller full-text sample size.

**Table III:** Comparative Analysis of GPT-3.5 Turbo’s Performance Metrics for Full-Text Inclusion Across Review Topics.

| Metric | <u>Dementia and Gait Signatures (10-fold)</u> |  | <u>Cuffless BP Monitoring (5-fold)</u> |  | <u>Long-Covid Outcomes (10-fold)</u> |  |
| --- | --- | --- | --- | --- | --- | --- |
|  | Control | Model | Control | Model | Control | Model |
| Accuracy | 0.53 ± 0.09 | 0.62 ± 0.10 | 0.41 ± 0.17 | 0.56 ± 0.21 | 0.47 ± 0.12 | 0.49 ± 0.07 |
| Sensitivity | 0.69 ± 0.27 | 0.69 ± 0.34 | 0.37 ± 0.42 | **0.95 ± 0.11** | 0.43 ± 0.42 | **0.89 ± 0.18** |
| Specificity | 0.52 ± 0.10 | 0.61 ± 0.12 | 0.40 ± 0.17 | 0.49 ± 0.24 | 0.47 ± 0.11 | 0.45 ± 0.07 |
| PPV | 0.14 ± 0.09 | 0.18 ± 0.12 | 0.14 ± 0.14 | 0.30 ± 0.15 | 0.08 ± 0.09 | 0.15 ± 0.08 |
| NPV | 0.92 ± 0.09 | 0.95 ± 0.04 | 0.77 ± 0.19 | 0.95 ± 0.11 | 0.87 ± 0.09 | **0.97 ± 0.05** |
| F1 Score | 0.22 ± 0.11 | 0.27 ± 0.16 | 0.20 ± 0.21 | 0.44 ± 0.17 | 0.14 ± 0.14 | 0.24 ± 0.12 |
| MCC | 0.10 ± 0.16 | 0.20 ± 0.18 | -0.16 ± 0.37 | **0.33 ± 0.22** | -0.07 ± 0.28 | **0.20 ± 0.12** |

Note: Metrics were derived using a pseudo-K-folds cross-validation method. BP – Blood Pressure, PPV – Positive Predictive Value, NPV – Negative Predictive Value, MCC – Matthews Correlation Coefficient. Bolded values indicate a p-value < .05.

### 3.7 Full-Text Self-Validation of AI Model Responses

Figure 4 extends our self-validation analysis to the full-text screening phase, presenting confusion matrices that compare the AI model’s responses when explanations were provided versus when they were not. Unlike the earlier phase, this figure visualizes outcomes for all processed full-text articles, including those rated as ‘included’ by either the human reviewer or the model during the initial title-abstract screening.

**Figure 4.**
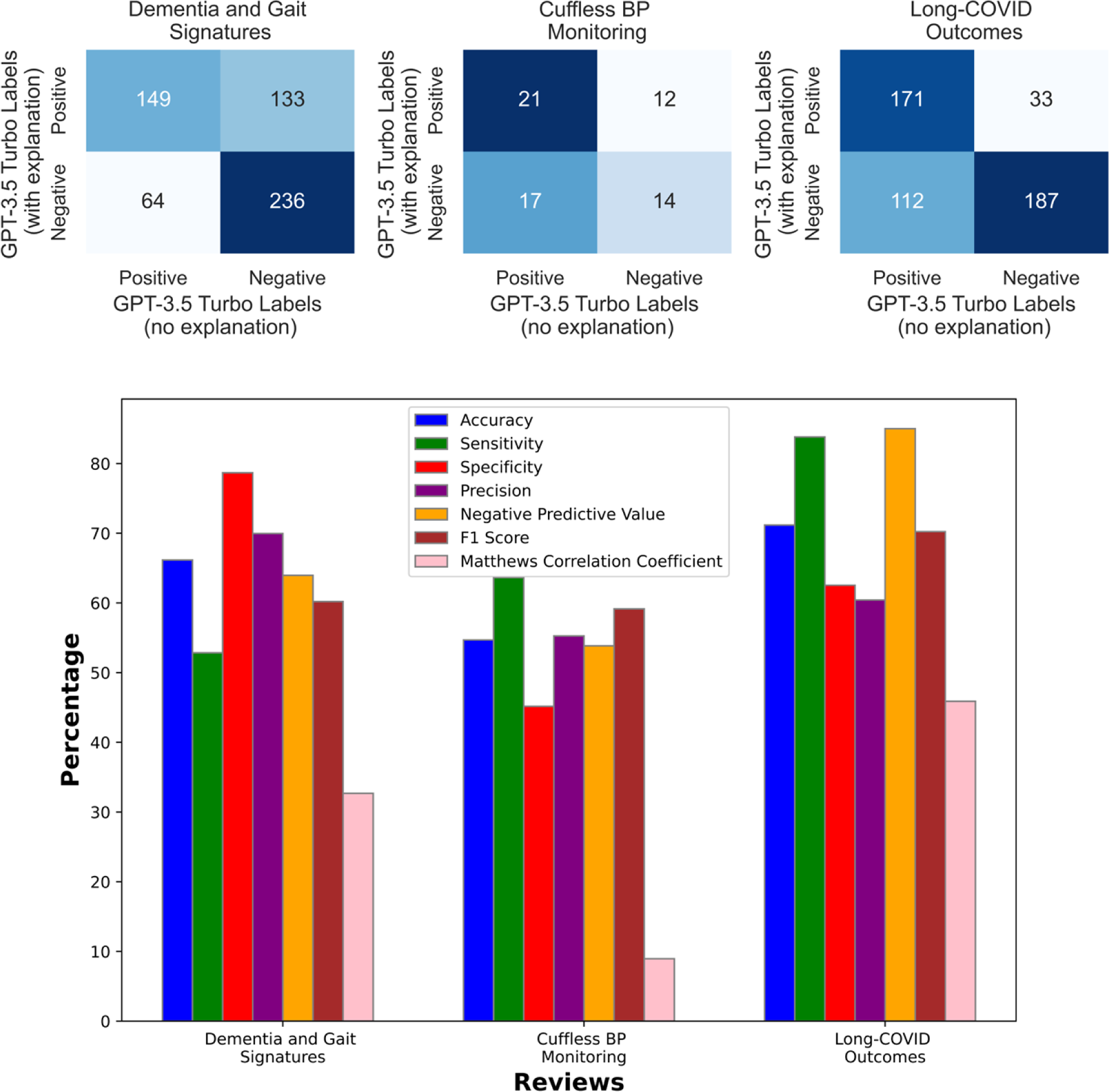
Comparative Analysis of GPT-3.5 Turbo Model Responses for Full-Text Screening: Evaluating Agreement Between Explanatory and Non-Explanatory Assessments Across Dementia Gait, Cuffless Blood Pressure Monitoring, and Long-COVID Outcomes Reviews. Confusion Matrices (top) and Performance Metrics (bottom).

The full-text phase shows lower levels of agreement between explanation and non-explanation modes. This diminished alignment may reflect variations in how effectively the text-embedding model retrieves relevant text snippets—which may require prompting and storage strategies separate from that of GPT-3.5 Turbo. It could also reflect differences in the model’s decision-making when evaluating more extensive article content, highlighting the need for further refinement in how AI models handle more complex data inputs.

## 4. Discussion

### 4.1 Role and Impact of AI and LLMs

The advent of AI and LLMs, epitomized by our LLM screening tool, marks a notable evolution in research methodologies for systematic reviews. LLMs such as BERT [20], Megatron-ML [25], GPT-3 [19], GPT-4 [23], and PaLM 2 [24], characterized by their large parameter sets and transformer architecture, have set new benchmarks in performance, illustrating the rapid evolution and potential of these models [6,22]. Our protocol facilitates a paradigm shift, enabling more efficient screening and precise extraction of information from the expansive realm of scientific literature. This advancement not only speeds up the discovery process but also enriches the insights gained from data [26], aligning with the growing need for swift review methodologies amid the surge of preprint repositories [6].

Further enriching our understanding, our model’s self-validation study offers a glimpse into the decision-making capabilities of AI. When comparing the model’s internally consistent responses, whether reasoned or not, against human reviewers’ decisions, a slight dip in performance was observed when the model explained its choices. This intriguing outcome hints at a complex, intuitive-like decision-making process within AI, akin to human cognition but distinct in its execution. These findings underscore the sophisticated nature of LLMs in systematic review processes and underscore the necessity for ongoing research to fully grasp AI’s potential in enhancing both the efficiency and depth of research synthesis.

### 4.2 Integration with Existing Methodological Innovations and Tools

The introduction of LLM screening into the ecosystem of systematic review tools such as PICO Portal, DistillerSR, Covidence, and Rayyan exemplifies a leap forward in the automation of review workflows [44–46]. These tools, alongside innovations like RobotReviewer [47], TrialStreamer [48], and Abstrackr [49], have showcased their capability in extracting and evaluating information from scientific articles, thus aiding in judging study quality, and inferring treatment effects.

Unlike traditional human review processes, which can be subject to subconscious biases and sometimes lack expert knowledge, LLMs provide a consistent and replicable framework for decision-making and have demonstrated proficiency across various fields. However, our preliminary findings suggest that the wording of prompts can significantly impact the representation of articles included by the model, as illustrated by self-validation tasks and performance metrics of similar prompts (see eFigure 1). As other studies have indicated, integrating such AI technologies poses challenges, particularly in balancing effective filtering with accurate identification of pertinent studies [50]. This delicate equilibrium is crucial as we further incorporate AI into systematic reviews, ensuring that we harness both AI capabilities and human expertise without compromising the integrity and depth of research synthesis.

### 4.3 Cautious Reliance on Automated Screening Systems

The deployment of open-source frameworks like LangChain [43] have demonstrated the transformative potential of AI and LLMs in enhancing various tasks, including the efficiency, accuracy, and overall workload involved in systematic reviews. By leveraging automation for the initial screening of papers, the tool has markedly reduced the time and manual labor traditionally required. Yet, amidst these advancements, we acknowledge the inherent limitations of AI systems in detecting nuanced or edge-case studies, a domain where human reviewers’ judgment and inclusivity play a crucial role.

The inclination of human reviewers to err on the side of inclusivity during the initial screening phases, adopting a ‘better safe than sorry’ approach to minimize the risk of overlooking potentially relevant studies, highlights a critical area where AI models may falter. GPT-3.5 Turbo, while adept at streamlining the review process, has exposed a vulnerability in terms of false negatives, underscoring the technology’s current limitations in fully grasping the subtleties of the full review procedure. This observation serves as a reminder of the need for cautious integration of AI-based screening systems within the systematic review workflow.

### 4.4. Data Validation and its Impact on Model Performance

Our evaluations have demonstrated GPT-3.5 Turbo’s effectiveness in managing heterogeneous datasets and identifying opportunities for iterative enhancement. Recent studies have highlighted the susceptibility of LLMs to the “butterfly effect,” where slight variations in input can precipitate significantly different outputs [51]. This sensitivity resonates with our findings from the self-validation tests, where subtle changes—such as requesting an explanation for decisions—markedly affected the model’s alignment between trials.

Given these findings, a critical focus of our analysis was the examination of data integrity and its influence on the AI model’s output. We observed minor inconsistencies in the reviewer-generated data, such as mismatches in titles, abstracts, and DOIs, which could potentially skew performance metrics. Manual restoration of these discrepancies had a negligible impact on the overall performance results, suggesting that the screening’s robustness extends to accommodating minor data inconsistencies.

### 4.5 Impact on Policy and Practice

While the adoption of AI, particularly LLMs like GPT-3.5 Turbo, offers transformative potential for systematic review methodologies, it introduces new vulnerabilities that warrant careful consideration. Central among these concerns is the danger posed by reliance on a single AI model for all decision-making processes within reviews. The risks of such a dependency include vulnerability to adversarial attacks, where manipulated inputs could lead to inaccurate outcomes, and the presence of inherent biases within the AI model that could skew results in subtle yet significant ways.

Addressing these risks involves not only technological solutions but also a broader reconsideration of how AI tools are integrated into the review ecosystem. It necessitates a balanced approach that leverages the strengths of AI for efficiency and scalability while maintaining a critical awareness of its limitations and potential pitfalls. As we advance, fostering a diversified toolkit of AI models and open-source methodologies will be paramount in mitigating the risks associated with overreliance on any single system. This strategy will enhance the resilience of the review process, ensuring that it remains robust, transparent, and adaptable to the evolving landscape of scientific inquiry.

During data collection, we encountered several practical challenges that underscore the current barriers to automating systematic review processes. Our study screening protocol was constrained by technical limitations, primarily due to reliance on textual inputs and difficulties handling multi-modal data, such as tables and figures. Additionally, even with access to institutional libraries, we faced obstacles in scraping for DOIs and accessing all relevant articles, hampered by publisher restrictions. These challenges highlight broader issues in the accessibility of scientific literature and point to the urgent need for infrastructural improvements. Such enhancements are crucial to support the seamless integration of AI tools in research synthesis and to ensure their effective utilization.

### 4.6 Future Directions

As we continue to refine our novel screening protocol, striking a balance between the speed and thoroughness of the review process remains a central challenge. Efforts are currently directed towards prompt engineering and comparing various models to minimize the exclusion of relevant literature. Future research should focus on enhancing the system’s ability to process multi-modal inputs and expanding its capabilities for comprehensive, end-to-end review automation. Such advancements could optimize research synthesis and enable LLMs to assimilate findings across multiple disciplines. This could lead to unique and innovative insights, potentially revolutionizing the processing and utilization of complex, interdisciplinary information.

As it stands, while we advocate for the use of AI in systematic reviews for its undeniable benefits, we also emphasize the importance of integrating these technologies judiciously, ensuring that they complement rather than replace the nuanced judgment of human reviewers. This approach aims to harness the strengths of both AI and human expertise, optimizing the systematic review process without compromising the integrity and depth of research synthesis.

## Conclusion

The development and application of our LLM screening protocol signifies a milestone in research methodology. In our comprehensive work involving the screening and validation of GPT-3.5 Turbo across three review topics and 24,534 studies, the model has demonstrated its potential as a sophisticated, efficient, and reliable model for systematic reviews and meta-analyses automation. Its ongoing evolution and refinement are paramount to keep pace with the rapid progression of scientific and technological innovations.

## Data Availability

All relevant data are within the manuscript and its Supporting Information files.

## Funding

M. Jamal, Deen, NSERC Discovery Grant (2018-2024)

Zheng, Rong NSERC CREATE for Smart Mobility for the Aging Population (2020-2026)

## Acknowledgements

We thank our peers and mentors for their guidance and insights throughout this research. Our gratitude also extends to the NSERC for their support. We appreciate the contributions of the academic community whose studies form the basis of this research article.

## Author Contributions

All listed authors have made substantial contributions to the conception, design, execution, or analysis of this work. Each author has participated sufficiently in the work to take public responsibility for appropriate portions of the content. Furthermore, the manuscript has been read and approved by all the listed authors, ensuring that questions related to the accuracy or integrity of any part of the work are appropriately investigated and resolved.

